# SNP rs7549881, near SGIP1 at 1p31.3, is significantly associated with digestive disorders and Parkinsonism in women

**DOI:** 10.1101/2024.10.04.24314910

**Authors:** Steven Lehrer, Peter Rheinstein

## Abstract

**Background:** A recent study identified a mutation in the SH3GL Interacting Endocytic Adaptor 1 (SGIP1) gene, located at 1p31.3, linked to early-onset Parkinsonism in 2 sisters in an Arab family with a history of young-onset Parkinson symptoms. SGIP1 had not been previously associated with Parkinsonism, a group of disorders including Parkinson’s disease (PD), characterized by motor dysfunction and cognitive decline. SGIP1 plays a role in endocytosis, particularly clathrin-mediated synaptic vesicle recycling, essential for synaptic proteostasis. In a fruit fly model, SGIP1 deletion resulted in synaptic dysfunction, impaired protein recycling, and brain cell degeneration, which mirrors Parkinsonism. We aimed to explore the genetic association of SGIP1 in Parkinson’s disease using data from the UK Biobank, specifically examining the nearby common intron variant rs7549881 at 1p31.3.

**Methods:** This study utilized data from the UK Biobank, a large biomedical database with genetic and health information from approximately 500,000 participants. We analyzed the association between rs7549881 and PD. The study included all participants with self-reported or ICD-diagnosed PD and restricted analysis to those with at least seven years of education. Genetic association analysis was performed using PLINK and SPSS, with logistic regression adjusting for covariates such as age, sex, and smoking. Additionally, a Phenome-Wide Association Study (PheWAS) was conducted to examine rs7549881 across a wide range of phenotypes.

**Results:** The study analyzed data from 385,629 subjects, with PD patients having a mean age of 63 ± 5 years. Among females, 26% of those homozygous for the rs7549881 GG allele had PD, compared to 22.2% who did not have PD (p = 0.004). This effect was not significant in males. Logistic regression revealed that male sex (OR 1.828, p < 0.001), age (OR 1.141 per year, p < 0.001), and constipation (OR 4.726, p = 0.001) increased PD risk, while cigarette smoking decreased it (OR 0.748, p < 0.001). The GG genotype was associated with increased PD risk when compared to the AA phenotype (OR 1.208, p = 0.015). PheWAS identified significant associations of rs7549881 with digestive disorders, including functional digestive disorders and non-infectious gastroenteritis.

**Conclusion:** The study demonstrated an association between the rs7549881 variant and PD in females, highlighting the potential role of SGIP1 in neurodegeneration. Additionally, the PheWAS findings link this variant to gastrointestinal disorders, supporting the emerging concept of a gut-brain axis in PD. The results suggest that SGIP1 may influence both neuronal and gastrointestinal functions, providing a new avenue for understanding the genetic mechanisms underlying Parkinson’s disease and its non-motor symptoms.

---

A recent study identified a mutation in the SH3GL Interacting Endocytic Adaptor 1 (SGIP1, 1p31.3) gene linked to early-onset Parkinsonism. This mutation was discovered in 2 sisters in an Arab family with a history of Parkinson’s symptoms that appeared at a young age. The SGIP1 gene had not been previously associated with Parkinsonism, a group of disorders characterized by motor dysfunction and cognitive decline, including Parkinson’s disease (PD). To study the effects of this mutation, researchers created a fruit fly model lacking SGIP1. These flies exhibited movement difficulties and degeneration in brain cells, mirroring Parkinsonism symptoms. The mutation caused defects in synapses. Specifically, the mutation impaired protein recycling at the synapse, disrupting synaptic proteostasis, which is crucial for maintaining healthy neurons [1].

The SGIP1 gene is involved in regulating endocytosis, a process critical for cellular function and synaptic transmission in neurons. It plays a role in clathrin-mediated endocytosis, which allows cells to internalize molecules like neurotransmitters and membrane receptors. *SGIP1* interacts with proteins involved in membrane dynamics and synaptic vesicle recycling, which are crucial for maintaining synaptic communication between neurons. Mutations in SGIP1 can disrupt these processes, leading to synaptic dysfunction, which has been linked to neurodegenerative diseases such as early-onset Parkinsonism. The proper function of SGIP1 is therefore essential for healthy neuronal signaling and synaptic plasticity [2].

In the current study, we used data from UK Biobank to assess the relationship between SGIP1 at 1p31.3 and PD by studying a nearby common polymorphism, rs7549881 at 1p31.3. We then performed a phenome wide association study (PheWAS) of rs7549881.

## Methods

The UK Biobank is a large-scale biomedical database and research resource containing in-depth genetic, lifestyle, and health information from around 500,000 participants. Launched in 2006, it is one of the most significant health studies in the world. The project collects data on various health measures such as blood pressure, physical activity, cognitive function, and genetic sequences, as well as information from medical records and biological samples (blood, urine, saliva).

The primary goal of the UK Biobank is to improve the prevention, diagnosis, and treatment of a wide range of serious and life-threatening illnesses, including cancer, heart disease, stroke, and dementia. Its data is available to approved researchers worldwide, making it a valuable resource for understanding the genetic and environmental factors that contribute to disease. The study’s vast scale allows researchers to explore patterns and correlations at an unprecedented level, leading to numerous discoveries in public health and medicine.

UK Biobank has approval from the Northwest Multi-center Research Ethics Committee (MREC) to obtain and disseminate data and samples from the participants, and these ethical regulations cover the work in this study. Written informed consent was obtained from all participants. Details can be found at www.ukbiobank.ac.uk/ethics.

Our UK Biobank study was approved as UKB 57245 (SL and PHR). Our analysis included all subjects with PD that occurred either before or after participant enrollment and was recorded in the UK Biobank database using self-reported data and the International Classification of Diseases (ICD10, ICD9). We restricted our analysis to subjects with at least 7 years of education, since subjects with less education could have problems with the touch screen self-entry data system.

We studied the relation to PD of SNP rs7549881,1p31.3, a common single nucleotide intron variant, A > G, minor allele frequency 0.43, 201 base pairs. Data processing was performed on Minerva, a Linux mainframe with Centos 7.6, at the Icahn School of Medicine at Mount Sinai. We used PLINK, a whole-genome association analysis toolset, to analyze the UKB chromosome files. Statistical analysis was done with SPSS 26.

We then performed PheWAS (Phenome-Wide Association Study), a method used to explore the associations between genetic variants (typically single nucleotide polymorphisms or SNPs) and a broad range of phenotypes or disease traits. Unlike the more familiar GWAS (Genome-Wide Association Study), which focuses on finding genetic variants linked to a specific trait or disease, PheWAS does the opposite: it starts with a genetic variant, in this case rs7549881, and then looks across multiple phenotypes to see what associations exist.

PheWAS helps identify new connections between genetic variants and various diseases or traits, potentially uncovering pleiotropy, where one genetic variant affects multiple outcomes. By leveraging large datasets, such as electronic health records (EHRs) or biobanks like the UK Biobank, PheWAS can analyze many phenotypes simultaneously, leading to insights into the genetic basis of complex diseases, drug repurposing opportunities, and personalized medicine. This approach has been instrumental in discovering unexpected links between genetic variants and conditions, further deepening our understanding of the genetic architecture of human health.

To perform PheWAS of rs7549881 we used the Phenome-Wide Association Study Browser PheWeb, a tool that allows researchers to explore the results of PheWAS analyses. PheWeb provides an interactive platform to visualize the associations between genetic variants (typically SNPs) and a wide array of phenotypes derived from large-scale biobank data, such as the UK Biobank and other genomic data resources.

## Results

Data from 385,629 subjects was analyzed. PD patients were aged 63 ± 5 (mean ± SD). 95% of subjects were white British.

Table 1 shows presence of Parkinson’s Disease versus rs7549881 genotype in female and male subjects. 26% of female subjects homozygous for the minor allele (GG) of rs7549881 had PD. 22.2% of female subjects homozygous for the minor allele (GG) of rs7549881 did not have PD. This difference was significant (p = 0.004, two tail Fisher exact test). The effect of rs7549881 genotype on male subjects was not significant (p = 0.183).

**Table 1.**
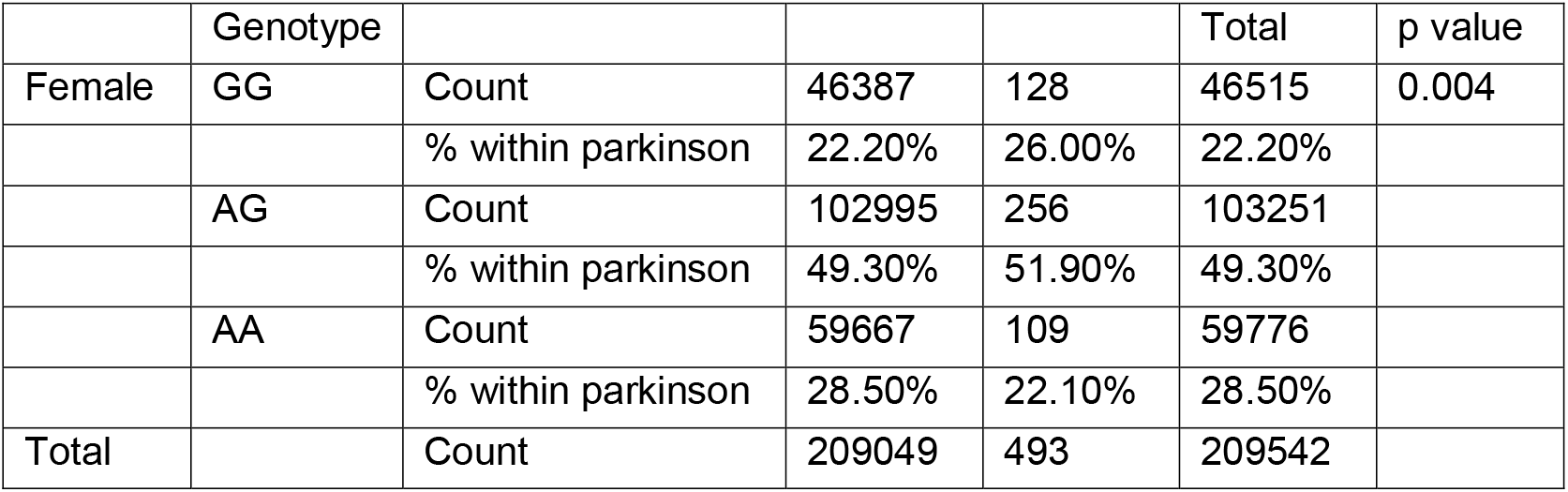
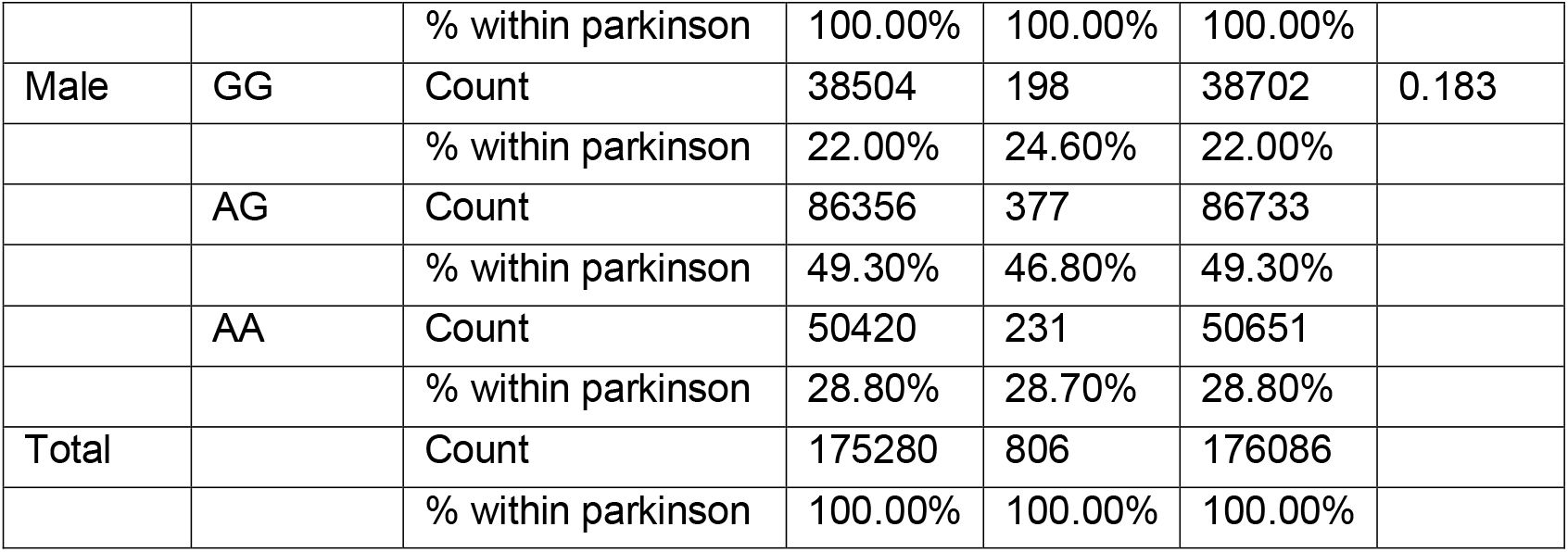
Presence of Parkinson’s Disease versus rs7549881 genotype in female and male subjects. 26% of female subjects homozygous for the minor allele (GG) of rs7549881 had PD. 22.2% of female subjects homozygous for the minor allele (GG) of rs7549881 did not have PD. This difference was significant (p = 0.004, two tail Fisher exact test). The effect of rs7549881 genotype on male subjects was not significant (p = 0.183).

Table 2 shows multivariate analysis of rs7549881/PD data with logistic regression. Male sex was associated with increased risk of PD (odds ratio O.R. 1.828, p < 0.001). Each year of age was associated with increased risk of PD (O.R. 1.141, p < 0.001). Cigarette smoking was associated with decreased risk of PD (O.R. 0.748, p < 0.001). Constipation was associated with increased risk of PD (O.R. 4.726, p = 0.001). rs7549881 GG genotype compared to AA genotype was associated with increased risk of PD (O.R. 1.208, p = 0.015).

**Table 2.** Multivariate analysis of PD data with logistic regression. Male sex was associated with increased risk of PD (odds ratio O.R. 1.828, p < 0.001). Each year of age was associated with increased risk of PD (O.R. 1.141, p < 0.001). Cigarette smoking was associated with decreased risk of PD (O.R. 0.748, p < 0.001). Constipation was associated with increased risk of PD (O.R. 4.726, p = 0.001). rs7549881 GG genotype compared to AA genotype was associated with increased risk of PD (O.R. 1.208, p = 0.015). (95% L.B. 95% confidence interval lower bound, U.B. 95% confidence interval upper bound).

|  | 95% L.B. | O.R. | 95% U.B. | p value |
| --- | --- | --- | --- | --- |
| sex | 1.634 | 1.828 | 2.044 | <0.001 |
| age | 1.130 | 1.141 | 1.152 | <0.001 |
| smoking | 0.647 | 0.748 | 0.865 | <0.001 |
| constipation | 1.933 | 4.726 | 11.551 | 0.001 |
| AA vs GG | 1.037 | 1.208 | 1.408 | 0.015 |
| GA vs GG | 0.927 | 1.057 | 1.205 | 0.417 |

Figure 1 shows PheWas of rs7549881. The most significant associations were functional digestive disorders, personal history of diseases of the digestive system, and non-infectious gastroenteritis.

**Figure 1.**
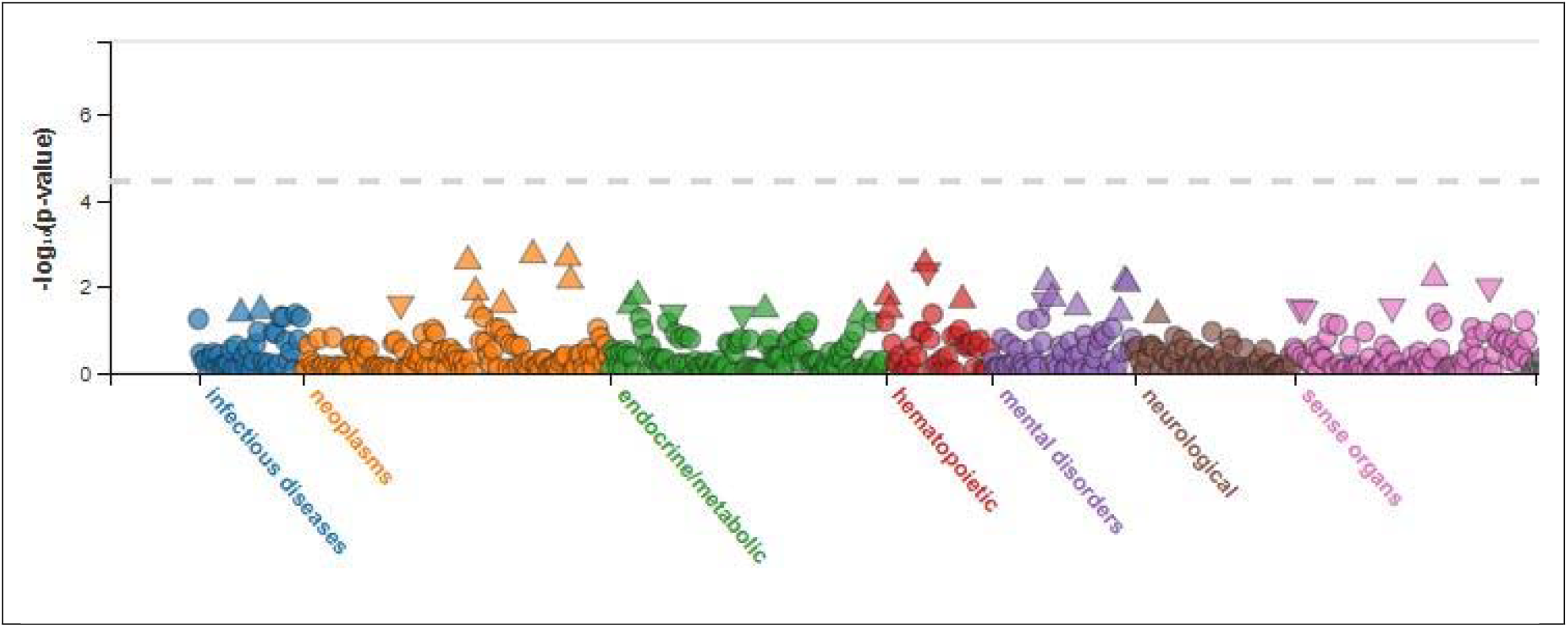
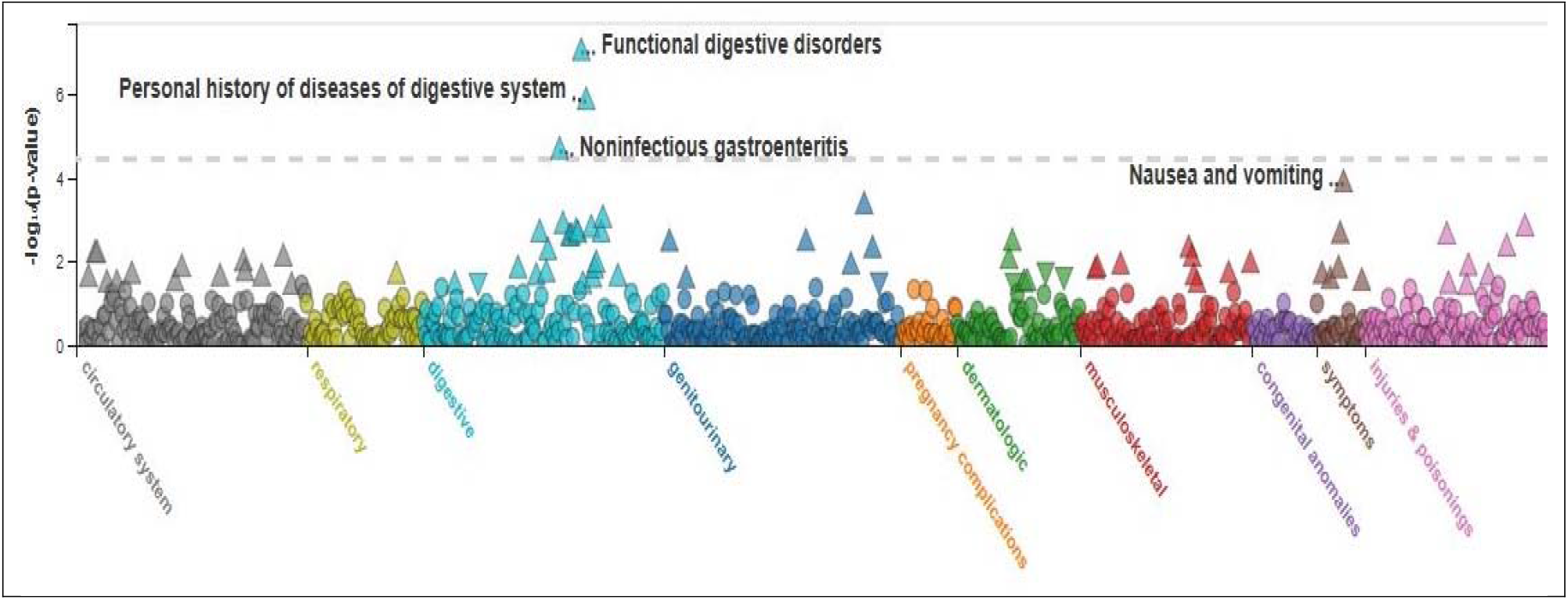
PheWAS of rs7549881. Note that the most significant associations were functional digestive disorders, personal history of diseases of the digestive system, and non-infectious gastroenteritis.

Table 3 shows ten most significant PheWas associations. The top 4 are digestive: functional digestive disorders, personal history of diseases of digestive system, noninfectious gastroenteritis, nausea and vomiting.

**Table 3.** Ten most significant rs7549881 PheWAS associations (se standard error). The top 4 are digestive: Functional digestive disorders, Personal history of diseases of digestive system, Noninfectious gastroenteritis, Nausea and vomiting.

| Category | Phenotype | P-value | Effect Size (se) | Number of samples (case/control) |
| --- | --- | --- | --- | --- |
| digestive | Functional digestive disorders | 1.00E-07 | 0.054 (0.010) | 21937 / 333366 |
| digestive | Personal history of diseases of digestive system | 1.50E-06 | 0.058 (0.012) | 15316 / 333367 |
| digestive | Noninfectious gastroenteritis | 2.30E-05 | 0.050 (0.012) | 15612 / 333376 |
| symptoms | Nausea and vomiting | 1.40E-04 | 0.051 (0.013) | 11651 / 395550 |
| genitourinary | Hypertrophy of female genital organs | 4.70E-04 | 0.14 (0.040) | 1268 / 207570 |
| digestive | Other chronic nonalcoholic liver disease | 1.00E-03 | 0.11 (0.035) | 1687 / 398277 |
| digestive | Intestinal obstruction without mention of hernia | 1.40E-03 | 0.073 (0.023) | 3939 / 333382 |
| injuries & poisonings | Symptoms concerning nutrition, metabolism, and development | 1.60E-03 | 0.062 (0.020) | 5488 / 401714 |
| digestive | Peritonitis and retroperitoneal infections | 1.70E-03 | 0.15 (0.047) | 927 / 385683 |
| neoplasms | Benign neoplasm of colon | 1.80E-03 | 0.033 (0.011) | 20121 / 384292 |

## Discussion

The SGIP1 gene encodes a protein that plays a crucial role in the process of endocytosis, particularly in neurons within the brain. Its primary functions are:

### Clathrin-Mediated Endocytosis

> SGIP1 is involved in clathrin-mediated endocytosis, a cellular process where cells internalize molecules (such as nutrients and neurotransmitters) by engulfing them in a clathrin-coated vesicle [3].
>
> SGIP1 interacts with proteins like Eps15 and endophilins, which are essential components of the endocytic machinery. These interactions facilitate the formation of clathrin-coated pits and vesicles [4].

### Synaptic Vesicle Recycling

> SGIP1 is highly expressed in the brain, especially in neurons. It is critical for the efficient recycling of synaptic vesicles—the tiny sacs that store neurotransmitters released during synaptic transmission. By aiding in the rapid retrieval and recycling of synaptic vesicle components after neurotransmitter release, SGIP1 ensures sustained and efficient synaptic communication between neurons [1].

### Regulation of Membrane Dynamics

> SGIP1 influences the curvature of cellular membranes, which is important for vesicle formation and trafficking. SGIP1 can bind to phosphoinositides, a type of lipid that plays a role in signaling pathways and membrane dynamics [4].

### Potential Implications in Disease are

#### Neurological Disorders

> Recent research has identified mutations in the SGIP1 gene that are potentially linked to early-onset Parkinsonism. These mutations may disrupt synaptic function, leading to neuronal degeneration. Given its role in synaptic vesicle recycling, alterations in SGIP1 function can lead to impaired synaptic transmission, which is a feature of several neurodegenerative diseases [1].

#### Neurodevelopmental Disorders

> Abnormalities in endocytic processes can affect neuronal development and connectivity, potentially contributing to developmental disorders [5].

The SGIP1 gene is essential for maintaining efficient neuronal communication through its role in endocytosis and synaptic vesicle recycling. By facilitating the internalization and recycling of synaptic components, SGIP1 supports the rapid and repeated firing of neurons necessary for normal brain function. Ongoing research continues to investigate its broader implications in neurological diseases and how targeting SGIP1 pathways might offer new therapeutic strategies.

Although SGIP1 has not been notably associated with the gastrointestinal (GI) tract, the connection of nearby SNP rs7549881 with functional digestive disorders is not surprising. Indeed, a recent genome-wide pleiotropic analysis demonstrated a role of the gut-brain axis in the shared genetic etiology between gastrointestinal tract diseases and psychiatric disorders [6]. This association is important because of the close relationship between digestive disorders and PD.

GI disorders are a common non-motor symptom of PD and are linked to the disease’s development and progression. Up to 80% of PD patients experience constipation, which usually begins before other symptoms. Additional common GI symptoms include bloating, nausea, vomiting, and gastroparesis. Some research suggests that the gut may be the initial site of pathological changes in PD. The accumulation of Lewy pathology in the enteric nervous system can lead to impaired motility and constipation [7]. Moreover, people with PD have reduced levels of some beneficial bacteria, like Prevotella, Faecalibacterium, and Roseburia, while others, like Bifidobacterium and Lactobacillus, are increased [8].

In conclusion, the SGIP1 gene at 1p31.3 plays a critical role in endocytosis and synaptic vesicle recycling, essential processes for maintaining healthy neuronal communication. The SGIP1mutation previously identified in 2 young sisters has provided new insights into its involvement in early-onset Parkinsonism, highlighting how synaptic dysfunction contributes to neurodegeneration [1]. Our study above demonstrates an association between nearby SNP rs7549881 at 1p31.3 and Parkinson’s disease, particularly in female subjects, as well as links to functional digestive disorders. These findings emphasize the potential for SGIP1 and rs7549881 to serve as therapeutic targets, not only in Parkinsonism but also in addressing the broader gut-brain axis, which may influence both neurological and gastrointestinal health. Ongoing research could lead to novel therapeutic approaches for neurodegenerative diseases.

## Data Availability

All data produced in the present study are available from PheWeb.org or from UK Biobank after approved study application

https://pheweb.org

## Acknowledgment

graphical abstract made with biorender.com

---

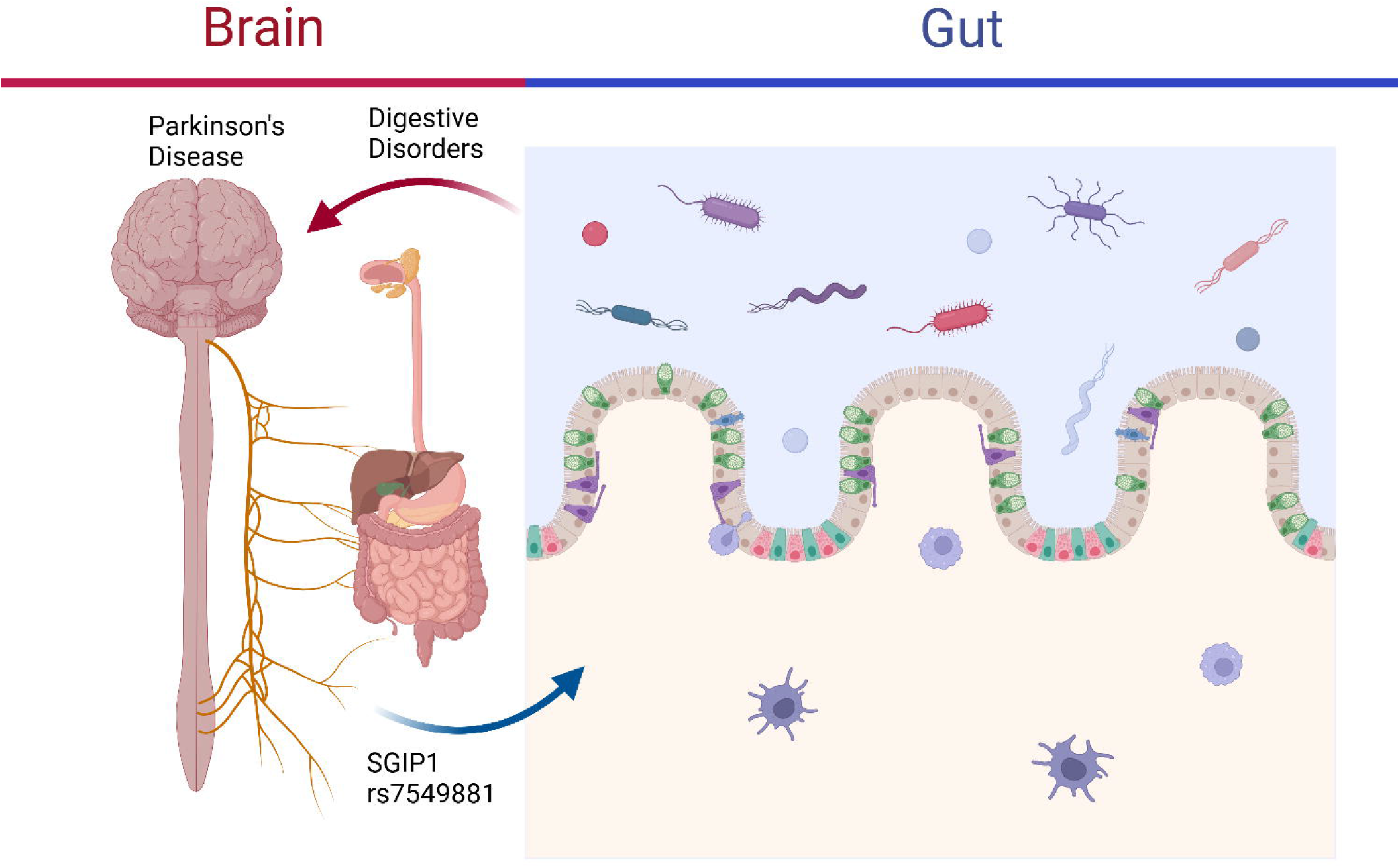

